# Time-Varying Mortality Risk Suggests Increased Impact of Thrombosis in Hospitalized Covid-19 Patients

**DOI:** 10.1101/2021.12.11.21267259

**Authors:** Benjamin J. Lengerich, Mark E. Nunnally, Yin Aphinyanaphongs, Rich Caruana

## Abstract

Treatment protocols, treatment availability, disease understanding, and viral characteristics have changed over the course of the Covid-19 pandemic; as a result, the risks associated with patient comorbidities and biomarkers have also changed. We add to the ongoing conversation regarding inflammation, hemostasis and vascular function in Covid-19 by performing a time-varying observational analysis of over 4000 patients hospitalized for Covid-19 in a New York City hospital system from March 2020 to August 2021 to elucidate the changing impact of thrombosis, inflammation, and other risk factors on in-hospital mortality. We find that the predictive power of biomarkers of thrombosis risk have increased over time, suggesting an opportunity for improved care by identifying and targeting therapies for patients with elevated thrombophilic propensity.

## Introduction

Treatment protocols, treatment availability, disease understanding, and viral characteristics have changed over the course of the Covid-19 pandemic; as a result, the risks associated with patient comorbidities and biomarkers have also changed. Analyses of hospitalized patients have identified that inflammatory biomarkers correspond to case severity [1, 2]; in addition, observations of clotting have suggested that hemostasis and vascular function are critically dysregulated processes in patients hospitalized with Covid-19 [3-8]. We add to this ongoing conversation by performing a time-varying observational analysis of over 4000 patients hospitalized for Covid-19 in a New York City hospital system from March 2020 to August 2021 to elucidate the changing impact of thrombosis, inflammation, and other risk factors on in-hospital mortality. We find that the predictive power of biomarkers of thrombosis risk have increased over time, suggesting an opportunity for improved care by identifying and targeting therapies for patients with elevated thrombophilic propensity. This strengthening of the association between thrombosis risk and mortality contrasts against a weakening association between biomarkers of inflammation risk and mortality.

## Methods

We use a generalized additive model (GAM) to predict in-hospital mortality from patient risk factors: demographics, vital signs, comorbidities, and initial lab tests on hospital admission. The GAM estimates a main effect of each risk factor and an interaction with time for each lab test. We use the GAM implemented in the Python InterpretML package [9] which is invariant to all monotonic feature transforms, and we estimate confidence intervals (CIs) by bootstrap resampling. To streamline analysis, we binarize each of the 11 continuous-valued lab tests into a discrete biomarker rule which approximately separates high-risk and low-risk regions (Table 1). For analysis, we group the 11 biomarkers into 3 groups: thrombosis risk (high D-Dimer, high hematocrit), inflammatory risk (high C-Reactive protein, high Neutrophil/Lymphocyte ratio, low serum albumin), and other biomarkers. The GAM achieves state-of-the-art accuracy for in-hospital mortality prediction (AUROC=0.933 on held-out patients) and provides odds ratios (ORs) for the risk associated with each biomarker on each day. We supplement these daily ORs with ORs from 3 logistic regression (LR) models each trained on approximately one third of the patients (LR1 was trained on patients hospitalized from day 1 to day 100, LR2 was trained on patients hospitalized from day 100 to 300, while LR3 was trained on patients hospitalized from day 300 to 527) and observe qualitatively similar patterns in the GAM and the LR models, although the GAM provides more statistical power and better temporal resolution.

**Table 1.**
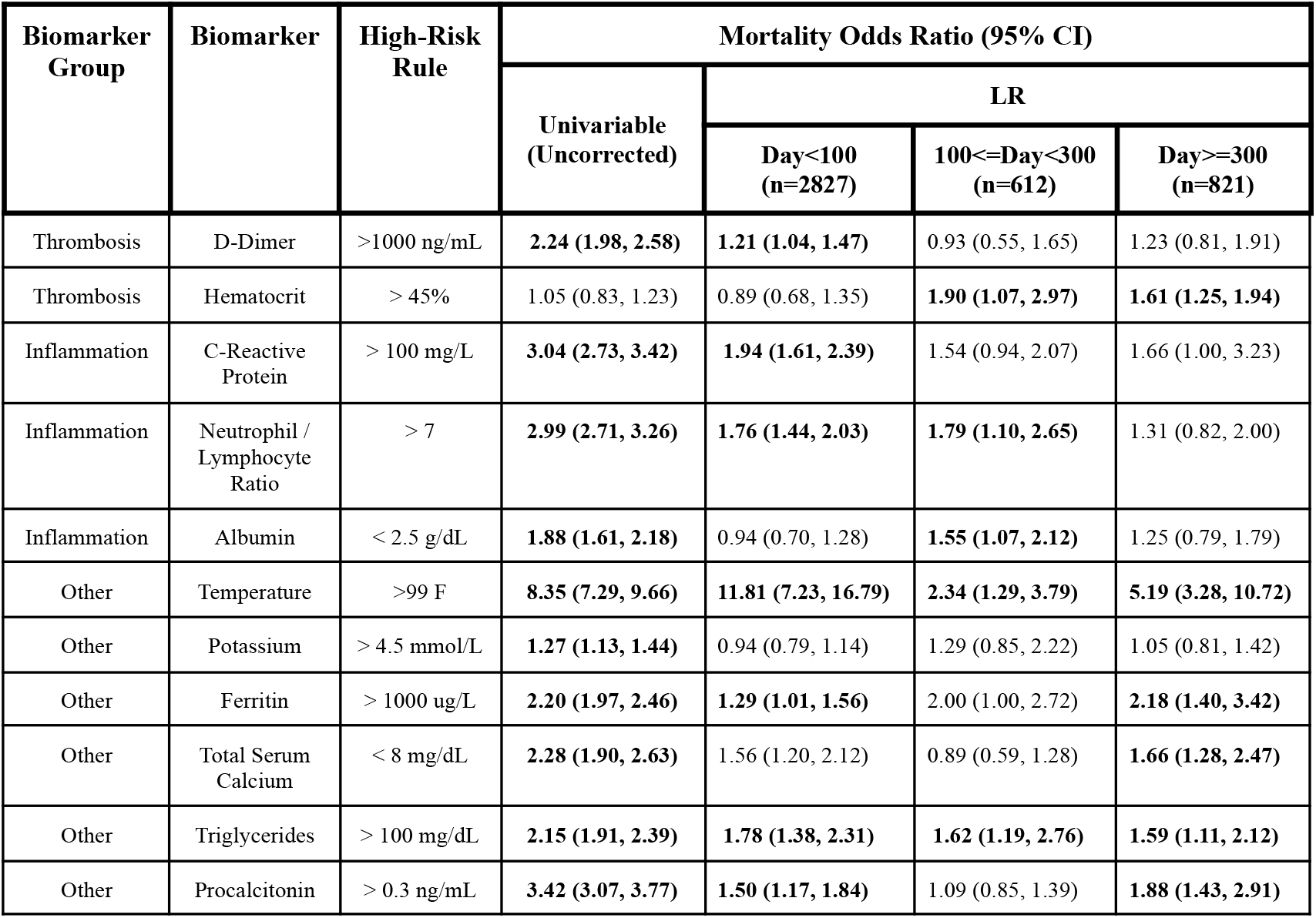
This Table lists the biomarkers and rules analyzed by the GAM presented in Figure 1. In addition, we supplement the statistically more powerful GAM results with odds ratios of in-hospital mortality under (1) a univariable analysis without any correction for confounding factors, (2) a multivariable logistic regression model trained on patients admitted in the first 100 days of the pandemic, (3) a multivariable logistic regression model trained on patients admitted from days 100 to 300 of the pandemic, and (4) a multivariable logistic regression model trained on patients admitted after day 300 of the pandemic. The strongest risk factor is elevated temperature, and the only risk factors estimated to consistently increase in predictive power under the logistic regression model are elevated ferritin and elevated hematocrit.

| Biomarker Group | Biomarker | High-Risk Rule | Mortality Odds Ratio (95% CI) |  |  |  |
| --- | --- | --- | --- | --- | --- | --- |
|  |  |  | Univariable (Uncorrected) | LR |  |  |
|  |  |  |  | Day<100 (n=2827) | 100<=Day<300 (n=612) | Day>=300 (n=821) |
| Thrombosis | D-Dimer | >1000 ng/mL | **2.24 (1.98, 2.58)** | **1.21 (1.04, 1.47)** | 0.93 (0.55, 1.65) | 1.23 (0.81, 1.91) |
| Thrombosis | Hematocrit | > 45% | 1.05 (0.83, 1.23) | 0.89 (0.68, 1.35) | **1.90 (1.07, 2.97)** | **1.61 (1.25, 1.94)** |
| Inflammation | C-Reactive Protein | > 100 mg/L | **3.04 (2.73, 3.42)** | **1.94 (1.61, 2.39)** | 1.54 (0.94, 2.07) | 1.66 (1.00, 3.23) |
| Inflammation | Neutrophil / Lymphocyte Ratio | > 7 | **2.99 (2.71, 3.26)** | **1.76 (1.44, 2.03)** | **1.79 (1.10, 2.65)** | 1.31 (0.82, 2.00) |
| Inflammation | Albumin | < 2.5 g/dL | **1.88 (1.61, 2.18)** | 0.94 (0.70, 1.28) | **1.55 (1.07, 2.12)** | 1.25 (0.79, 1.79) |
| Other | Temperature | >99 F | **8.35 (7.29, 9.66)** | **11.81 (7.23, 16.79)** | **2.34 (1.29, 3.79)** | **5.19 (3.28, 10.72)** |
| Other | Potassium | > 4.5 mmol/L | **1.27 (1.13, 1.44)** | 0.94 (0.79, 1.14) | 1.29 (0.85, 2.22) | 1.05 (0.81, 1.42) |
| Other | Ferritin | > 1000 ug/L | **2.20 (1.97, 2.46)** | **1.29 (1.01, 1.56)** | 2.00 (1.00, 2.72) | **2.18 (1.40, 3.42)** |
| Other | Total Serum Calcium | < 8 mg/dL | **2.28 (1.90, 2.63)** | 1.56 (1.20, 2.12) | 0.89 (0.59, 1.28) | **1.66 (1.28, 2.47)** |
| Other | Triglycerides | > 100 mg/dL | **2.15 (1.91, 2.39)** | **1.78 (1.38, 2.31)** | **1.62 (1.19, 2.76)** | **1.59 (1.11, 2.12)** |
| Other | Procalcitonin | > 0.3 ng/mL | **3.42 (3.07, 3.77)** | **1.50 (1.17, 1.84)** | 1.09 (0.85, 1.39) | **1.88 (1.43, 2.91)** |

## Cohort

Our dataset consists of patients hospitalized in the NYU Langone Health system who have lab-confirmed cases of Covid-19. To filter out patients who were hospitalized for reasons other than Covid-19, we excluded patients who have indicators of (1) pregnancy: outpatient prenatal vitamins, in-patient oxytocics, folic acid preparations; (2) scheduled surgery: urinary tract radiopaque diagnostics, laxatives, general anesthetics, antiemetic/antivertigo agents, or (3) parasitic infection: in-patient nitazoxanide. We also require that the patients have recorded temperature, age, BMI, admission day, and Neutrophil-Lymphocyte ratio. Finally, we remove patients who died within six hours of admission. After this filtering, we have 4260 patients.

For each patient, the model includes demographics, comorbidities, outpatient medications, initial in-patient vitals, and initial in-patient lab tests (Supplement S1). To remove confounding with end-of-life care, we exclude measurements taken within 24 hours of mortality. The highest density of patients were admitted in April 2020 (days 30-50 of the pandemic); this period also contained many patients with high underlying risk (Figure S1).

## Results

For each day, we estimate the contribution to mortality risk from each biomarker, and the mean mortality risk contributed by the thrombosis-related biomarkers and inflammation-related biomarkers after correcting for confounding from all other risk factors. The association between thrombosis biomarkers and in-hospital mortality has strengthened over time, rising from an OR of 0.92 (95% CI 0.85-1.00) in March 2020 to an OR of 1.55 (1.38-1.69) in August 2021 (Figure 1A). This rise in thrombosis risk contrasts against a decrease in risk associated with inflammation-related biomarkers, which has dropped from an OR of 1.42 (1.37-1.49) in March 2020 to 1.16 (1.03-1.28) in August 2021. While other biomarkers have had stable impacts on mortality risk (e.g. the risk associated with elevated temperature has remained consistently strong), the risk associated with elevated ferritin has increased from an OR of 1.29 (1.07-1.49) to 2.22 (1.82-2.69) in August 2021. These trends are qualitatively corroborated by LR models (Table 1).

**Figure 1.**
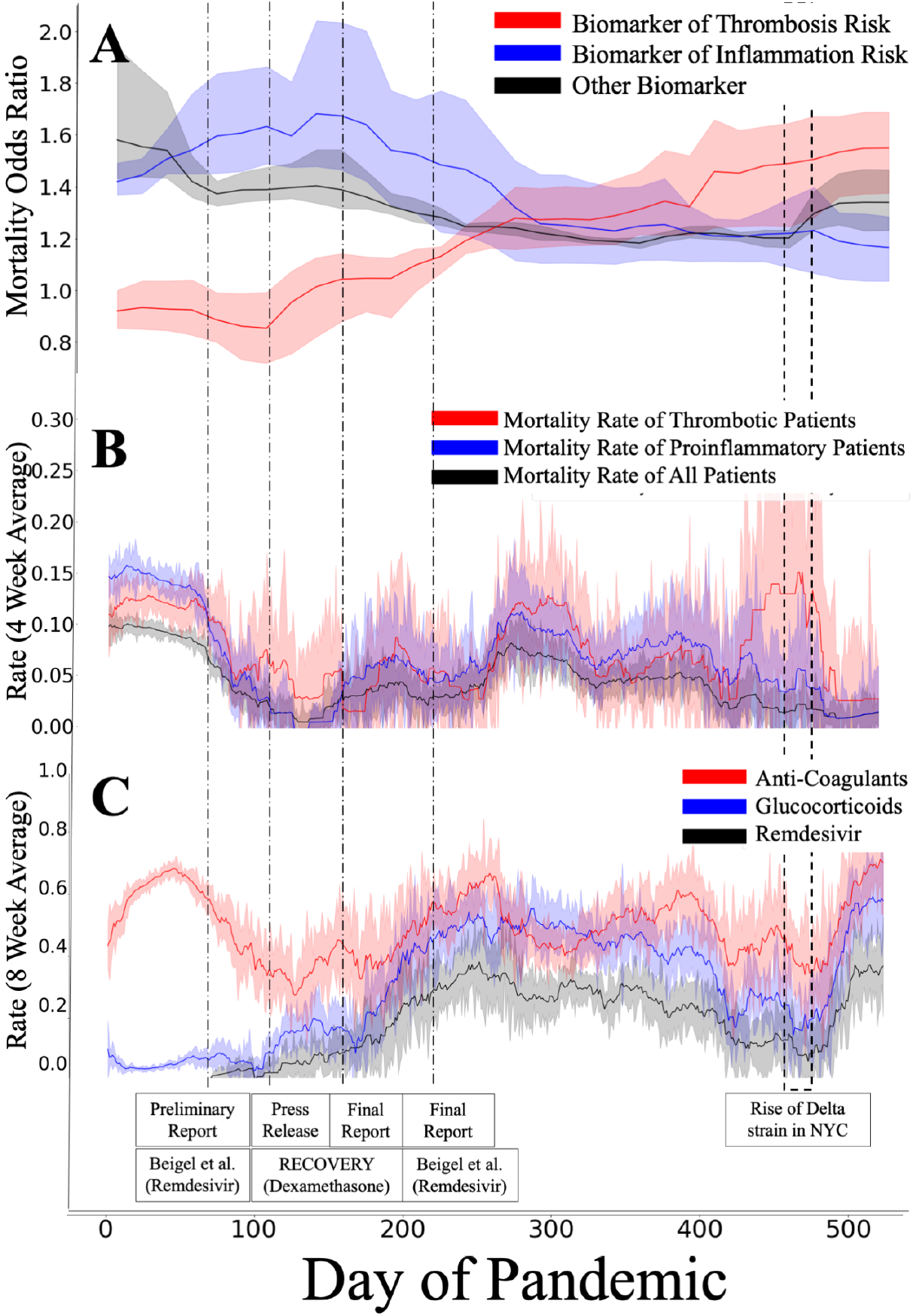
(**A)** The predictive power of biomarkers have changed over time. Biomarkers of inflammation risk (elevated C-reactive protein, low albumin, high Neutrophil/Lymphocyte ratio) were initially powerful predictors of in-hospital mortality, but have become less predictive over time. In contrast, biomarkers of thrombosis risk (elevated D-Dimer, elevated hematocrit) are more predictive of mortality in August 2021 than during March 2020. This suggests that the successful treatment of patients hospitalized with indicators of thrombosis risk has lagged behind the treatment of other groups. **(B)** The in-hospital mortality rate has decreased over time for all patients, but at a reduced rate for patients satisfying at least one biomarker rule for thrombosis risk. **(C)** Treatment protocols have changed over time, with a trend toward glucocorticoid and anticoagulant prescription (overwhelmingly prophylactic heparin) for the majority of patients. We mark the dates of several important publications and the rise of the Delta strain in NYC.

These trends are also weakly observed even in raw mortality rates: the overall mortality rate has decreased from 20.3% (16.6%-24.1%) in March 2020 to 3.5% (0.8%-6.2%) in August 2021, and the mortality rate of patients with at least 1 biomarker of inflammatory risk has similarly decreased from 29.9% (25.7%-34.3%) in March 2020 to 3.7% (0.0%-12.6%) in August 2021, while the mortality rate of patients with at least 1 biomarker of thrombosis risk has only decreased from 22.6% (14.3%-30.9%) in March 2020 to 6.3% (0.0%-13.5%) in August 2021 (Figure 1B). Finally, these trends in mortality risk partially correspond to trends in prescription rates (Figure 1C): clinical trials have suggested the utility of glucocorticoids [10] and remdesivir [11], and the prescription rates of these treatments increased following publication of these studies. However, despite recognition of the importance of thrombosis and clotting in Covid-19 [3-8], anticoagulant prescription rates have varied as the effectiveness of thromboprophylaxis with heparin has been questioned [12,13].

## Discussion

These results suggest that success in care for patients at risk for thrombosis has lagged behind the success in care for patients with biomarkers of inflammation. As with all observational analyses, this study has limitations. Notably, without randomizing interventions, it is difficult to identify a singular cause driving these trends. Several hypotheses would be consistent with these data, including: (1) the SARS-CoV-2 Delta strain shifted the importance of intrinsic risk factors, (2) successful efforts for early detection and treatment of thrombosis risk [16] widened the difference in mortality risk between a serologically defined prothrombotic state and active thromboses, (3) potential interactions between anti-inflammation treatments and haemostasis, vascular function, and thrombophilic propensity [14,15], or (4) a lack of effective thromboprophylaxis treatments in Covid-19: there is little evidence for thromboprophylaxis from heparin in Covid-19 patients [12,13] which may be due to heparin’s reliance on endogenous Antithrombin (AT) [17] which can be reduced in Covid-19 patients [18] --- anticoagulants such as Argatroban [19] or Bivalirudin [20] which do not rely on AT may exert more powerful thromboprophylaxis than heparin in Covid-19 patients, or (5) an alternate process linked to thrombosis risk factors but also potentially implicating other aspects of endothelial and vascular dysfunction. All in all, our results suggest that it may be beneficial to focus more research on patients who have thrombosis or are at high thrombotic risk.

## Data Availability

The data include anonymized medical records, which are not publicly available.

## Acknowledgements

Yin Aphinyanaphongs was partially supported by NIH 3UL1TR001445-05 and National Science Foundation award #1928614.

The authors declare no relevant conflicts of interest.

## Author Contributions

BL, YA, and RC conceived the study design. BL implemented the statistical analysis. BL, MN, YA, and RC analyzed results and wrote the manuscript.

## Notes

### Competing Interest Statement

The authors have declared no competing interest.

### Author Declarations

The NYU Langone Health Institutional Review Board (IRB) determined that the proposed activity is not research involving human subjects. IRB review and approval is not required.

