## Supplement for "Time-Varying Mortality Risk Suggests Increased Impact of Thrombosis in Hospitalized Covid-19 Patients"

#### S1 Cohort Description

Table S1 presents the distribution in the studied cohort of the 11 biomarkers listed in Table 1 of the main text.

| Risk Factor | Mean Value | 25th Percentile | 75th Percentile | Missing Rate (%) |
|---|---|---|---|---|
| Albumin | 3.28 | 3.00 | 3.08 | 3.5 |
| C-Reactive Protein (mg/l) | 84.21 | 15.18 | 131.30 | 11 |
| Calcium | 8.68 | 8.30 | 9.10 | 0.19 |
| D-dimer (ng/ml) | 802.53 | 158.00 | 647.00 | 18.95 |
| Ferritin | 910.56 | 140.03 | 1084.72 | 12.58 |
| Hematocrit | 37.61 | 34.00 | 41.80 | 0 |
| Neutrophil/Lymphocyte Rate | 6.74 | 2.95 | 8.70 | 0 |
| Potassium (mmol/l) | 4.13 | 3.70 | 4.50 | 0.24 |
| Procalcitonin | 1.06 | 0.02 | 0.25 | 23.05 |
| Triglycerides | 63.8 | 0.00 | 109.00 | 55.06 |
| Temperature (F) | 98.29 | 97.80 | 98.60 | 0 |

**Table S1.** The distribution of biomarkers in the observed cohort.

Tables S2 and S3 present the risk factors used to correct for underlying patient risk.

| Risk Factor | Mean Value | 25th Percentile | 75th Percentile | Missing Rate (%) |
|---|---|---|---|---|
| Age (Years) | 64.36 | 53 | 78 | 0 |
| Bmi (kg / m^2) | 25.18 | 22.13 | 31.16 | 0 |
| Cancer (any malignancy) | 0.17 | 0 | 0 | 0 |
| Cerebrovascular Disease | 0.26 | 0 | 1 | 0 |
| Charlson Score | 2.84 | 1 | 4 | 0 |
| Chronic Obstructive Pulmonary Disease | 0.32 | 0 | 1 | 0 |
| Congestive Heart Failure | 0.25 | 0 | 0 | 0 |
| Day | 128.27 | 29 | 233 | 0.02 |
| Dementia | 0.15 | 0 | 0 | 0 |
| Diabetes With Chronic Complications | 0.23 | 0 | 0 | 0 |
| Diabetes Without Chronic Complications | 0.43 | 0 | 1 | 0 |
| Hemiplegia Or Paraplegia | 0.05 | 0 | 0 | 0 |
| Metastatic Solid Tumour | 0.06 | 0 | 0 | 0 |
| Mild Liver Disease | 0.15 | 0 | 0 | 0 |
| Moderate Or Severe Liver Disease | 0.03 | 0 | 0 | 0 |
| Myocardial Infarction | 0.32 | 0 | 1 | 0 |
| Peptic Ulcer Disease | 0.04 | 0 | 0 | 0 |
| Peripheral Vascular Disease | 0.29 | 0 | 1 | 0 |
| Race | 3.38 | 2 | 5 | 0 |
| Renal Disease | 0.33 | 0 | 1 | 0 |
| Male | 0.6 | 0 | 1 | 0 |
| Afibrillation | 0.33 | 0 | 1 | 0 |
| Gastroesophageal reflux disease | 0.05 | 0 | 0 | 0 |
| Hypertension | 0.22 | 0 | 0 | 0 |
| Rheumatoid Arthritis | 0.37 | 0 | 1 | 0 |
| Sjorgen's syndrome | 0.03 | 0 | 0 | 0 |
| Valve Disease | 0.03 | 0 | 0 | 0 |
| Valve Replacement | 0.16 | 0 | 0 | 0 |

**Table S2.** The distribution of comorbidities and demographic risk factors in the studied cohort.

| Risk Factor | Mean Value | 25th Percentile | 75th Percentile | Missing Rate (%) |
|---|---|---|---|---|
| Analgesic/antipyretics,non-salicylate | 0.03 | 0 | 0 | 0 |
| Anticonvulsants | 0.03 | 0 | 0 | 0 |
| Antihyperglycemic, Biguanide Type | 0.03 | 0 | 0 | 0 |
| Statins | 0.1 | 0 | 0 | 0 |
| Benign Prostatic Hypertrophy/micturition Agents | 0.03 | 0 | 0 | 0 |
| Beta-Adrenergic Agents, Inhaled | 0.05 | 0 | 0 | 0 |
| Beta-adrenergic Blocking Agents | 0.04 | 0 | 0 | 0 |
| Blood Sugar Diagnostics | 0.03 | 0 | 0 | 0 |
| Calcium Channel Blocking Agents | 0.04 | 0 | 0 | 0 |
| Laxatives And Cathartics | 0.04 | 0 | 0 | 0 |
| Loop Diuretics | 0.03 | 0 | 0 | 0 |
| Platelet Aggregation Inhibitors | 0.07 | 0 | 0 | 0 |
| Proton-pump Inhibitors | 0.04 | 0 | 0 | 0 |
| Vitamin D Preparations | 0.05 | 0 | 0 | 0 |

**Table S3.** The distribution of outpatient medications taken before hospitalization (limited to medication classes taken by at least 100 patients). These are also used as risk factors to correct for underlying patient risk.

### S2 Extended Results

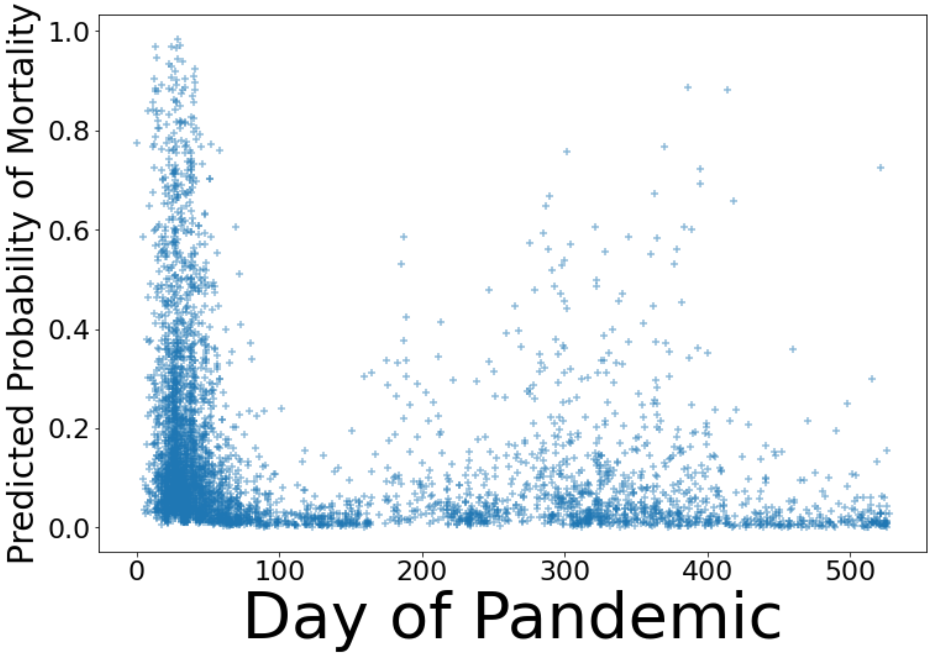


**Figure S1.** Predicted probability of mortality and admission day for each patient. In this figure, each point represents an individual patient, with the vertical location indicating the probability of mortality predicted by the mortality risk model. There is a high density of patients in the first 60 days of the pandemic, and the most high-risk patients were also observed at that time.

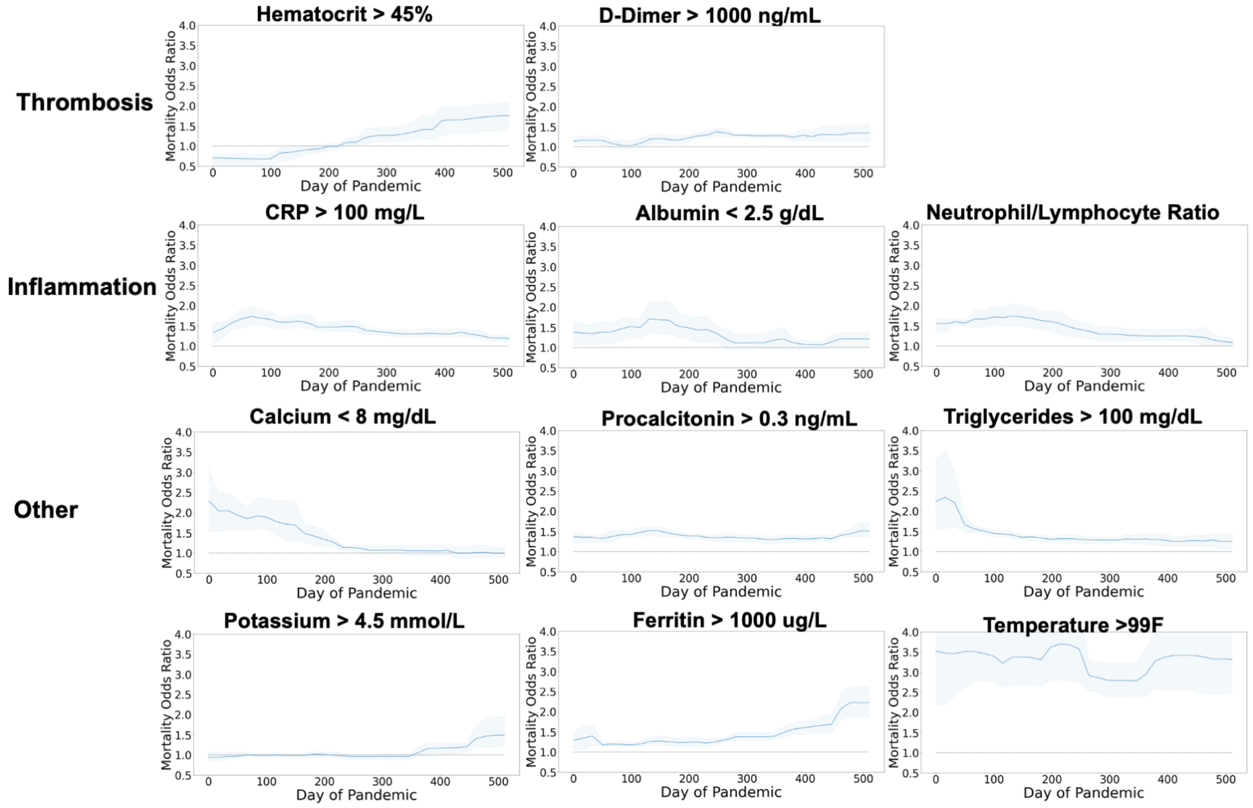


**Figure S2.** Effects of all biomarkers over the course of the pandemic. These effects are estimated by the GAM after correcting for all other confounding effects underlying patient risk. These effects are the same as those presented in Figure 1A of the main text.

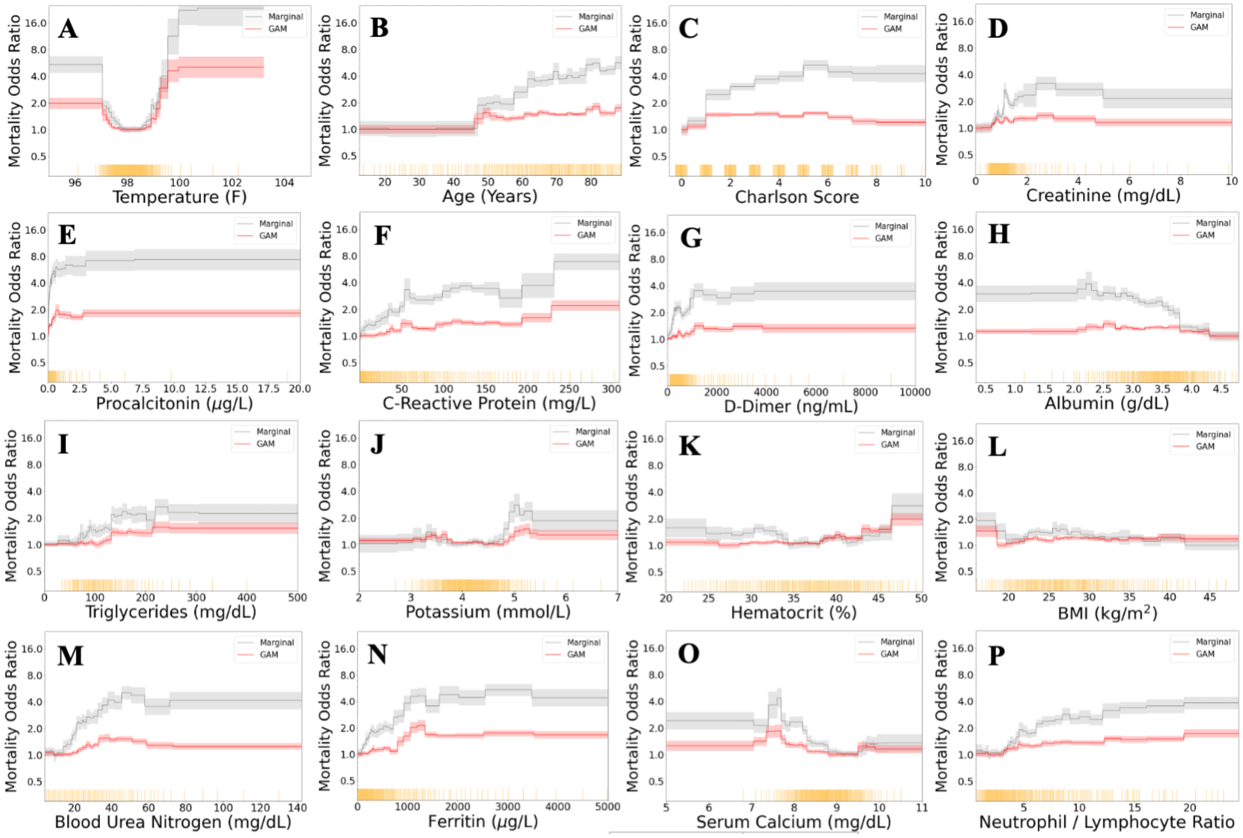


**Figure S3.** Main (not time-varying) effects of continuous-valued lab tests and vital signs. In each pane, we plot the effect estimated by the GAM which corrects for other confounding factors (red), and the effect estimated by marginalization without any correction (gray). Each yellow tick mark along the horizontal axis denotes 10 patients.
